# Estimation of country-level basic reproductive ratios for novel Coronavirus (COVID-19) using synthetic contact matrices

**DOI:** 10.1101/2020.02.26.20028167

**Authors:** Joe Hilton, Matt J. Keeling

## Abstract

The outbreak of novel coronavirus (COVID-19) has the potential for global spread, infecting large numbers in all countries. In this case, estimating the country-specific basic reproductive ratio is a vital first step in public-health planning. The basic reproductive ratio (*R*_0_) is determined by both the nature of pathogen and the network of contacts through which the disease can spread - with this network determined by socio-demographics including age-structure and household composition. Here we focus on the age-structured transmission within the population, using data from China to inform age-dependent susceptibility and synthetic age-mixing matrices to inform the contact network. This allows us to determine the country-specific basic reproductive ratio as a multiplicative scaling of the value from China. We predict that *R*_0_ will be highest across Eastern Europe and Japan, and lowest across Africa, Central America and South-Western Asia. This pattern is largely driven by the ratio of children to older adults in each country and the observed propensity of clinical cases in the elderly.

## 1 Introduction

The ongoing outbreak of 2019 novel coronavirus (2019-nCoV) has been characterised by a pattern of spread with most cases occur in older individuals, and in particular very few cases have been seen in under-15s[1, 2]. This suggests that transmission is characterised by age-specific heterogeneities going beyond those explained by differences in contact patterns across age groups.

A 2017 study by Prem *et al*. estimated contact patterns for 152 countries based on social and demographic data. These are in the form of matrices whose entries correspond to the expected total number of age-stratified contacts per day for individuals belonging to each of 16 5-year age classes[3]. These estimated contact matrices are publicly available as supplementary material attached to Prem *et al*.’s paper. Here we use the estimated social contact matrix for China to define the early age-structured transmission dynamics; this is formed from the country-specific social contact matrix ***k*** and a vector of age-specific susceptibilities *<u>z</u>* – whose entries scale the risk of transmission from contact with infected individuals to susceptible individuals of each age class. Based on age-structured data from the outbreak in China, we can estimate *<u>z</u>*. We then combine *<u>z</u>* with Prem *et al*.’s estimates of the contact matrices in the other 151 countries to generate an estimate of an age-structured transmission structure for each of those countries. The basic reproductive ratio can then be estimated from these transmission structures to give us an approximation of the spreading potential of a COVID-19 outbreak in each of these countries.

## 2 Methods

Our estimation process consists of calculating an age-specific susceptibility profile based on epidemiological data from China and the estimated China-level contact matrix[3]. This susceptibility profile can then be combined with the estimated country-level contact matrices from the other countries in Prem *et al*.’s study[3] to produce age-stratified transmission matrices for these countries. The dominant eigenvalue from these transmission matrices (which are linear scalings of the next-generation matrix) provides a relative estimate of the basic reproductive ratio for that country.

We assume that we have a population divided into *K* discrete age classes *C*_1_, …*C*_*K*_. Suppose we have a set of outbreak data in the form *<u>x</u>* = (*x*_1_, …, *x*_*K*_), where *x*_*i*_ is the cumulative number of cases so far in age class *C*_*i*_, expressed as a fraction of all the cases, so that ∑_*i*_ *x*_*i*_ = 1. Denote by *k*_*i,j*_ the expected number of contacts with individuals of age class *C*_*i*_ made per day by a single individual of age class *C*_*j*_. The matrix ***K*** = (*k*_*i,j*_) will be asymmetric, with the *i*th row corresponding to an average individual of each age class’s contacts with age class *C*_*i*_, and the *i*th column corresponding to the contacts made per day by an average individual of age class *C*_*i*_. Let *p*_*i*_ be the conditional probability that a susceptible individual in age class *C*_*i*_ becomes infected, given that they have had contact with an infectious individual. We assume that this probability depends only on the age class of the recipient, and not on that of the infector, i.e. we have age-dependent susceptibility but homogeneous infectivity. The expected number of infections generated in age class *C*_*i*_ in a single day by an infectious indvidual in age class *C*_*j*_ is given by *p*_*i*_*k*_*i,j*_. Defining *z*_*i*_ = *p*_*i*_*γ*^−1^, the expected number of cases in age class *C*_*i*_ generated by a single infectious individual in age class *C*_*j*_ over its entire infectious period is given by

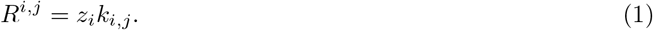

These age-stratified reproductive ratios define a matrix ***R*** (often called the next generation matrix), whose leading eigenvalue is the basic reproductive ratio of the entire system[4]. The corresponding magnitude 1 eigenvector is the distribution of cases by age in the early stages of the epidemic. Thus, given the dataset *<u>x</u>* and an estimate 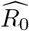 of the associated basic reproductive ratio, the matrix ***R*** should satisfy

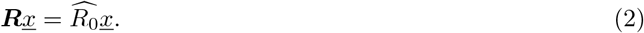

If we look at the *i*th row of this set of equations, we have

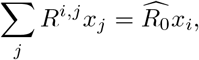

or, given Equation 1,

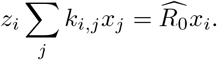

The age-specific susceptibility *z*_*i*_ can thus be estimated as

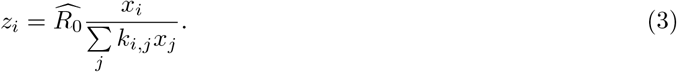

To estimate the equivalent of the matrix ***R*** for some new population with age-structured contact matrix 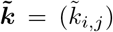, we assume that *z*_*i*_ is constant across populations (i.e. in every population we see the same dependence of susceptibility on age). Then our desired matrix has (*i, j*)th entry

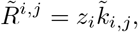

and the eigenvalue of this matrix gives us the estimated basic reproductive ratio 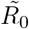 for the new population.

Since Equation 3 tells us that every *z*_*i*_ term is linear in 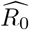, and every entry of 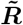 contains one of the *z*_*i*_ terms, it follows that the eigenvalue 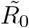 is also linear in 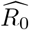. We can thus write 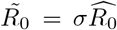 for some population-specific scaling factor *σ*, which we can calculate by carrying out the estimation of *<u>z</u>* with 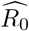 set to one. We can therefore calculate a full set of scaling factors for every country without requiring an estimate of the basic reproductive ratio in China; these scaling factors provide a measure of transmission relative to that observed in China, and can be used to generate country-specific basic reproductive ratios given a reliable estimate for China. Although we note that estimates for China differ widely depending on the time-scales examined (*R*_0_ = 2.2 [1], *R*_0_ = 2.3 − 2.6 [5], *R*_0_ = 2 − 2.7, [6], *R*_0_ = 2.35 [7], *R*_0_ = 3.11 [8]).

## 3 Results

In Figure 1 we present the results of a null-model in which there is no age-specific susceptibility (*z*_*i*_1), and plot the associated scaling factor *σ* for each of the 152 countries included in Prem *et al*.’s study. That is, we assume that the reproduction matrix ***R*** is directly proportional to the contact matrix ***K***. The constant of proportionality will be the ratio between the leading eigenvalues of the two matrices, i.e. *R*_0_ divided by the leading eigenvalue of ***K***. To obtain the reproduction matrix for some other country, we just multiply its contact matrix by the constant of proportionality we estimate for China.

**Figure 1:**
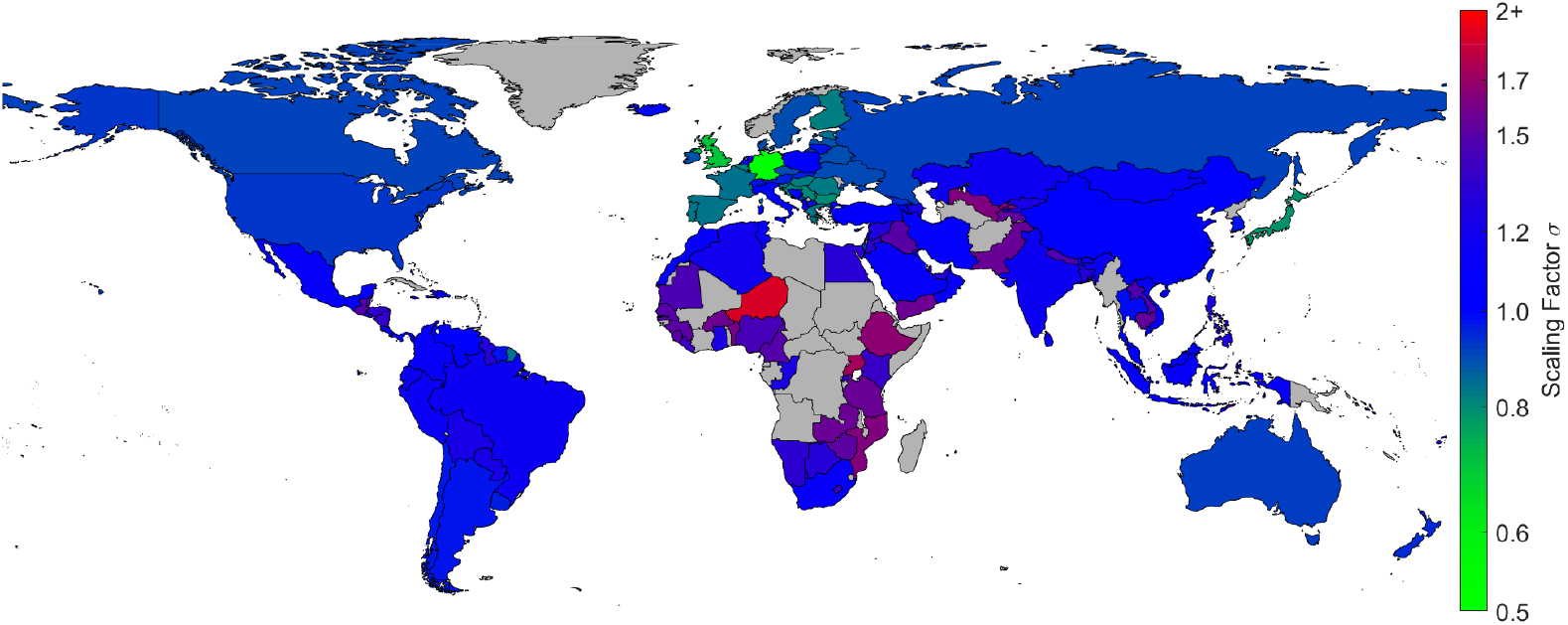
Estimated scaling factor *σ* for each country assuming no age-specific susceptibility. Gray countries are those not included in Prem *et al*.’s study [3].

Figure 2 presents the estimated scaling factors based on the age distribution reported by two studies of the early dynamics in China: Li *et al*.[1] which reports on the first 425 confirmed cases; and Yang *et al*.[2] which examines data from the first 4021 confirmed cases. (Our estimates of the scaling factors for each of the 152 countries is given in the Supplementary Material.)

**Figure 2:**
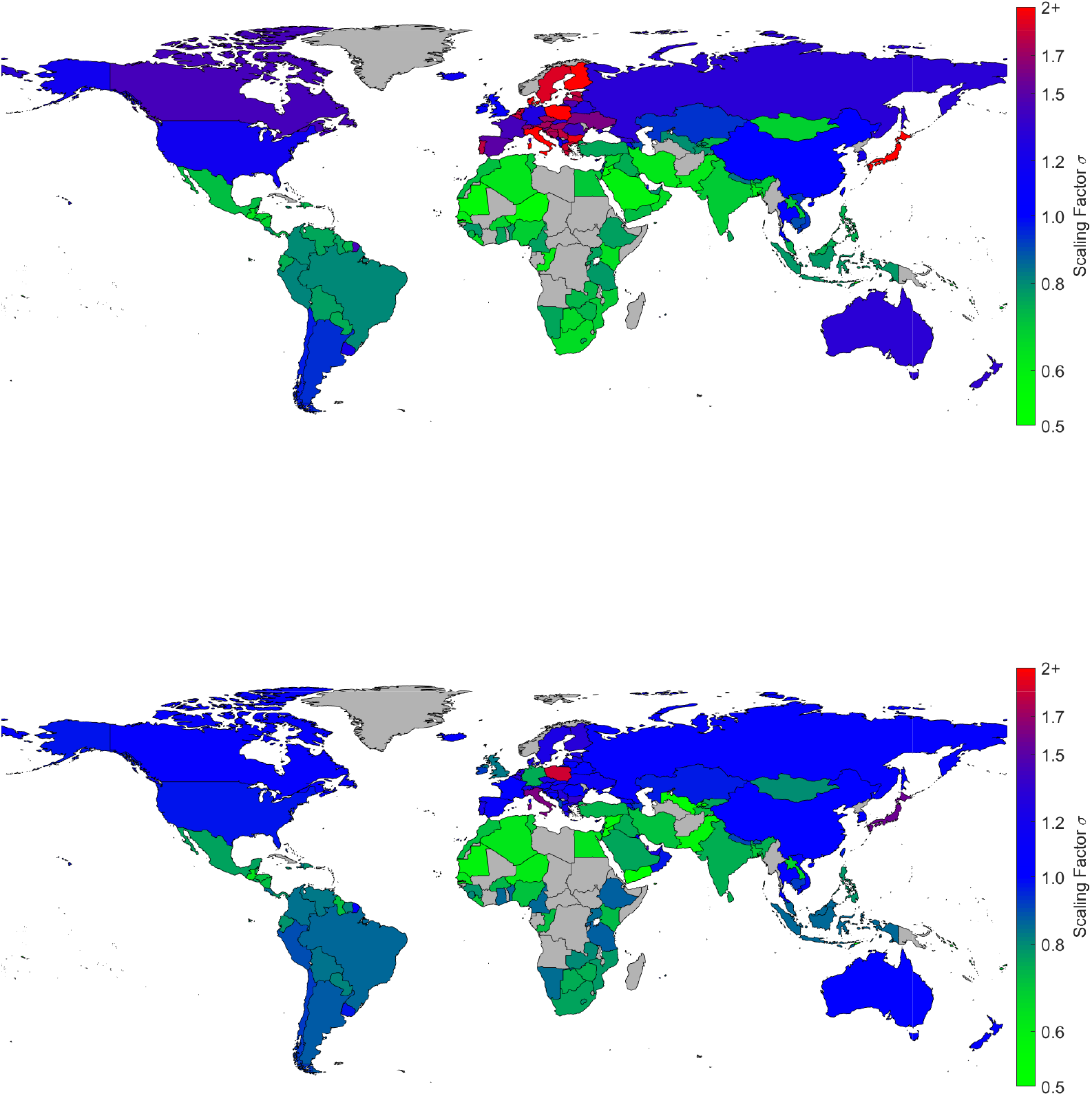
Estimated scaling factor *σ* for each country based on case data from Li *et al*. [1] (upper map) and Yang *et al*.[2] (lower map). Gray countries are those not included in Prem *et al*.’s study [3].

In Figure 3(a) we plot the scaling factors obtained through our consideration of age-dependent susceptibility (figure 2) against the factors obtained by assuming homogeneous susceptibility across age groups (figure 1). Allowing for age-dependent susceptibility substantially increases the amount of variation in scaling (and thus in basic reproductive ratio) by country, and in particular can lead us to predict much larger scaling factors. When the transmission structure is modelled as a simple scaling of a population’s contact patterns, the variation in scaling is driven by the variation in average intensity of contacts (captured by the eigenvalue of the contact pattern matrix ***K***) by country. When we incorporate age-dependent susceptibility, contact patterns involving members of highly susceptible age classes becomes particularly important - generating a core-group within the population. For COVID-19, this means that we see higher scaling factors in countries with older populations, since this usually means contacts involving older individuals are more common. In countries with comparitively younger populations, contacts involving older individuals are less common and so the capacity of the infection to spread is reduced relative to the purely-contact pattern based transmission model.

**Figure 3:**
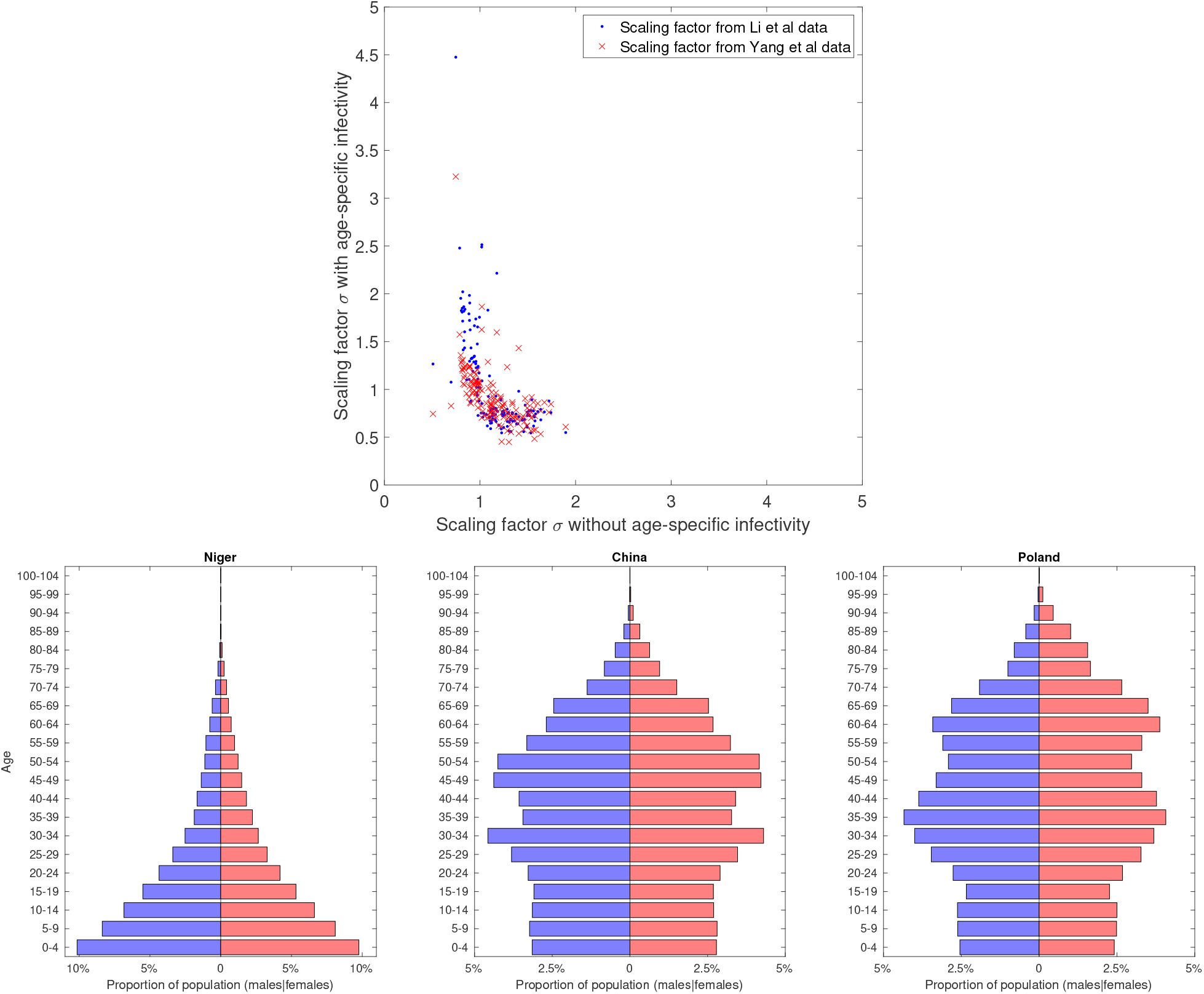
(a) Scaling factor estimated using outbreak data and the age-dependent susceptibility model versus scaling factor estimated without age-dependent susceptibility, based on age-dependent susceptibility profiles estimated from the Li *et al*. data (blue dots) and the Yang *et al*. data (red crosses). (b) Population pyramids for Niger, China and Poland - which have the highest, medium and lowest scaling respectively. Data from [9].

Figure 3(b) illustrates this principle for Niger (which is predicted to have one of the lowest scalings of the reproductive ratio) and Poland (which is predicted to have one of the highest scalings) in comparison to China. The population pryamid of Niger is dominated by young children; China has a relatively stable age-structure although there are more individuals in 30-54 age classes than in younger age-groups; the pryamid for Poland shows even fewer children and substantial proportions into older age-classes. We therefore observe that it is interaction between the population pryamid and the age-structured susceptiblity that largely drives the scaling of the basic reproductive ration.

This point is seen to hold for all countries investigated (Figures 1 and 2). In Figure 1, we see that without age-dependent susceptibility transmission is low in many European countries as well as in South Korea and Japan, and high in many African countries, consistent with the differences in daily number of contacts predicted by Prem *et al* driven by the proportion of children. However, the maps in Figure 2 demonstrate that when age-specific susceptibility is taken into account, the pattern of infectious potential by country is generally reversed. We then expect to see higher transmission in Eastern Europe (including Italy which had the largest number of cases in Europe in late February 2020) and Japan, and reduced transmission across Africa, central America, the Middle East and India.

## 4 Discussion

Here we have developed a simple model for age-structured transmission of 2019-nCoV with two components: an age-structured contact matrix dependent on the behaviour of the host population and an age-dependent susceptibility profile dependent on physiological response to infection. By using a previously-estimated synthetic contact matrix and age-stratified data, we were able to estimate age-dependent susceptibility profiles based on the first 425 and the first 4,021 cases in China. We then combined these estimated profiles with estimates of age-stratified contacts in 151 other countries to give us transmission matrices for these countries from which we could estimate the scale of basic reproductive ratios in each country relative to China. We demonstrated that taking age-specific susceptibility into account results in substantially different predictions of transmission intensity by country relative to a model without age-specific susceptibility; countries with older populations are at substantially higher risk than countries with younger populations.

The predictions we have made are limited by two main elements. The first is the accuracy of the estimated contact matrices; although there are known issues (as discussed in [3]) they remain our best estimate of age-structured contacts to date. Unfortunately, not all countries have an associated mixing matrix; many counties in Africa do not have the underlying demographic data to support the generation of the associated mixing matrix. Secondly, in inferring the age-dependent susceptibility, we are effectively generating a matrix *R*^*i,j*^ which determines the distribution of secondary confirmed cases in terms of current confirmed cases. We are therefore assuming that either younger individuals are unlikely to infected or if infected they are generally asymptomatic and play a minor role in onward transmission. If such asymptomatic infections transmit equally to symptomatic cases, then the scaling of the reproductive ratio is expected to be closer to Figure 1. Finally, it is worth stressing that these projections only inform about early phase of the outbreak in the absence of controls; the rapid and effective use of non-pharmaceutical interventions (contact-tracing, self-isolation and movement controls) can substantially reduce the reproductive ratio.

## Data Availability

The work only publically available data.

## 5 Acknowledgements

This research was funded by: the National Institute for Health Research (NIHR) (project reference 17/63/82) using UK aid from the UK Government to support global health research JH & MJK; the Engineering and Physical Sciences Research Council (EPSRC) (grant reference EP/S022244/1) MJK; Health Data Research UK, which receives its funding from HDR UK Ltd (NIWA1) MJK. The views expressed in this publication are those of the author(s) and not necessarily those of the any of the funders.

